# The Natural History of Vision-Related Quality of Life after Unilateral Occipital Stroke

**DOI:** 10.1101/2023.11.29.23299210

**Authors:** Neil Dogra, Bryan V. Redmond, Selena Lilley, Brent A. Johnson, Byron L. Lam, Madhura Tamhankar, Steven E. Feldon, Berkeley Fahrenthold, Jingyi Yang, Krystel R. Huxlin, Matthew R. Cavanaugh

## Abstract

**Background/Objectives:** Stroke damage to the primary visual cortex induces large, homonymous visual field defects that impair daily living. Here, we asked if vision-related quality of life (V-QoL) is impacted by time since stroke.

**Subjects/Methods:** We conducted a retrospective meta-analysis of 95 occipital stroke patients (F/M=26/69, 27-78 years old, 0.5 to 373.5 months post-stroke) in whom V-QoL was estimated using the National Eye Institute Visual Functioning Questionnaire (NEI-VFQ) and its 10-item neuro-ophthalmic supplement (Neuro10). Visual deficit severity was represented by the Perimetric Mean Deviation (PMD) calculated from 24-2 Humphrey visual fields. Data were compared with published cohorts of visually-intact controls. The relationship between V-QoL and time post-stroke was assessed across participants, adjusting for deficit severity and age with a multiple linear regression analysis.

**Results:** Occipital stroke patients had significantly lower NEI-VFQ and Neuro10 composite scores than controls. All subscale scores describing specific aspects of visual ability and functioning were impaired except for ocular pain and general health, which did not differ significantly from controls. Surprisingly, visual deficit severity was not correlated with either composite score, both of which increased with time post-stroke, even when adjusting for PMD and age.

**Conclusions:** V-QoL appears to improve with time post-occipital stroke, irrespective of visual deficit size or patient age at insult. This may reflect the natural development of compensatory strategies and lifestyle adjustments. Thus, future studies examining the impact of rehabilitation on daily living in this patient population should consider the possibility that their V-QoL may change gradually over time, even without therapeutic intervention.

## Introduction

Occipital stroke is the leading source of damage to the human primary visual cortex (V1; Zhang, Kedar, Lynn, Newman, & Biousse, 2006), causing a loss of conscious vision over similar portions of the visual field through both eyes (Gilhotra, Mitchell, Healey, Cumming, & Currie, 2002; Pollock et al., 2012; Pollock et al., 2011). This condition is known by many names but in this manuscript, it will be referred to as cortically-induced blindness (CB). It affects a significant portion of stroke survivors, with an estimated 1% of the population over age 49 years likely to develop CB in their lifetime (Gilhotra et al., 2002), and ∼100,000 new cases each year in the US and Europe (Gray et al., 1989; Pollock et al., 2012; Pollock et al., 2011; Rowe, 2013; Sahraie, 2007).

Activities of daily living including reading, driving, navigation, and autonomy are severely impaired in CB (Dombovy, Sandok, & Basford, 1986; Jones & Shinton, 2006; Jongbloed, 1986). Until recently, this vision loss was considered irreversible, with most patients discharged without rehabilitation opportunities (Horton, 2005a, 2005b; Plant, 2005; Pollock et al., 2019; Reinhard et al., 2005). As a result, there is little follow-up or monitoring of progression in CB, despite its significant impact on visual quality of life (V-QoL). However, accumulating experimental evidence suggests that visual training can recover a range of perceptual abilities within CB fields (for review, see Liu, Hanly, Fahey, Fong, & Bye, 2019; Melnick, Tadin, & Huxlin, 2016; Saionz, Busza, & Huxlin, 2022; Saionz, Feldon, & Huxlin, 2020). As restoration therapies targeting CB continue to develop, a better understanding of the natural evolution of V-QoL and the major factors driving these changes is essential to correctly interpret the impact of both the condition, and its rehabilitation, on daily living.

The National Eye Institute Visual Functioning Questionnaire (NEI-VFQ) is a clinically validated, self-administered survey that has been extensively used to evaluate V-QoL in CB patients (Chen et al., 2009; Gall, Franke, & Sabel, 2010; Gall, Lucklum, Sabel, & Franke, 2009; George, Hayes, Chen, & Crotty, 2011; Papageorgiou et al., 2007; Rowe et al., 2019). It asks respondents to assess difficulties they face with vision-specific functioning in contexts that include social gatherings, workplace performance, and pursuit of personal hobbies (Mangione, et al., 2001; Mangione et al., 1998). Responses from the questionnaire are used to assign numeric scores to 12 subscales: general health, general vision, ocular pain, near activities, distance activities, social functioning, mental health, role difficulties, dependency, driving, color vision, and peripheral vision. A composite score describing overall V-QoL is then generated by averaging all subscale scores except for general health. The NEI-VFQ can also be administered with a 10-item neuro-ophthalmic supplement (Neuro10), which generates an independent, composite score describing neuro-ophthalmic functions (Raphael et al., 2006). All scores scale from 0 to 100, with higher scores indicating better functioning.

One prior study found lower NEI-VFQ composite scores in 33 patients with stroke-induced V1 damage compared to a control group of healthy subjects (Papageorgiou et al., 2007). The authors reported that time since stroke and visual deficit severity did not affect NEI-VFQ scores, but only patients in the chronic phase (≥6 months) post-stroke were included (Papageorgiou et al., 2007). Similar findings were reported in another sample of 177 first-ever, chronic stroke patients (Gall et al., 2010). Finally, a study that included both acute and sub-acute patients (n=66) reported a large impact of deficit severity on V-QoL, but did not assess the effect of time-post stroke (Tharaldsen et al., 2020). Here, we examined V-QoL in 95 CB patients with V1 damage from a single etiology (occipital stroke), and in the absence of any therapeutic intervention for their vision loss. Our goal was to assess if V-QoL differed for patients <1 month to several years post-stroke. We also gauged the relative influence of deficit size/severity and participant age on this natural history. Finally, by examining individual subscale scores, we asked which aspects of visual or social functioning drove observed differences.

## Methods

### Participants

We conducted a retrospective, meta-analysis of V-QoL data from 95 patients with homonymous vision loss from stroke-induced damage to the occipital cortex, confirmed by magnetic resonance imaging or computed tomography (**Fig. 1A**). Patients were enrolled in one of two vision restoration clinical trials (ClinicalTrials.gov identifiers: NCT04798924, NCT03350919), or in experimental studies conducted by the Huxlin laboratory (NCT05098236). Exclusion criteria in all studies were: best-corrected visual acuity worse than 20/40, concurrent use of medications that might affect test performance, presence of ocular or neurologic condition(s) other than occipital stroke, which might cause visual impairment or impede study performance. By including data from 3 different trials, we were able to include a larger number of patients and times post-stroke than were present in individual studies.

**Figure 1.**
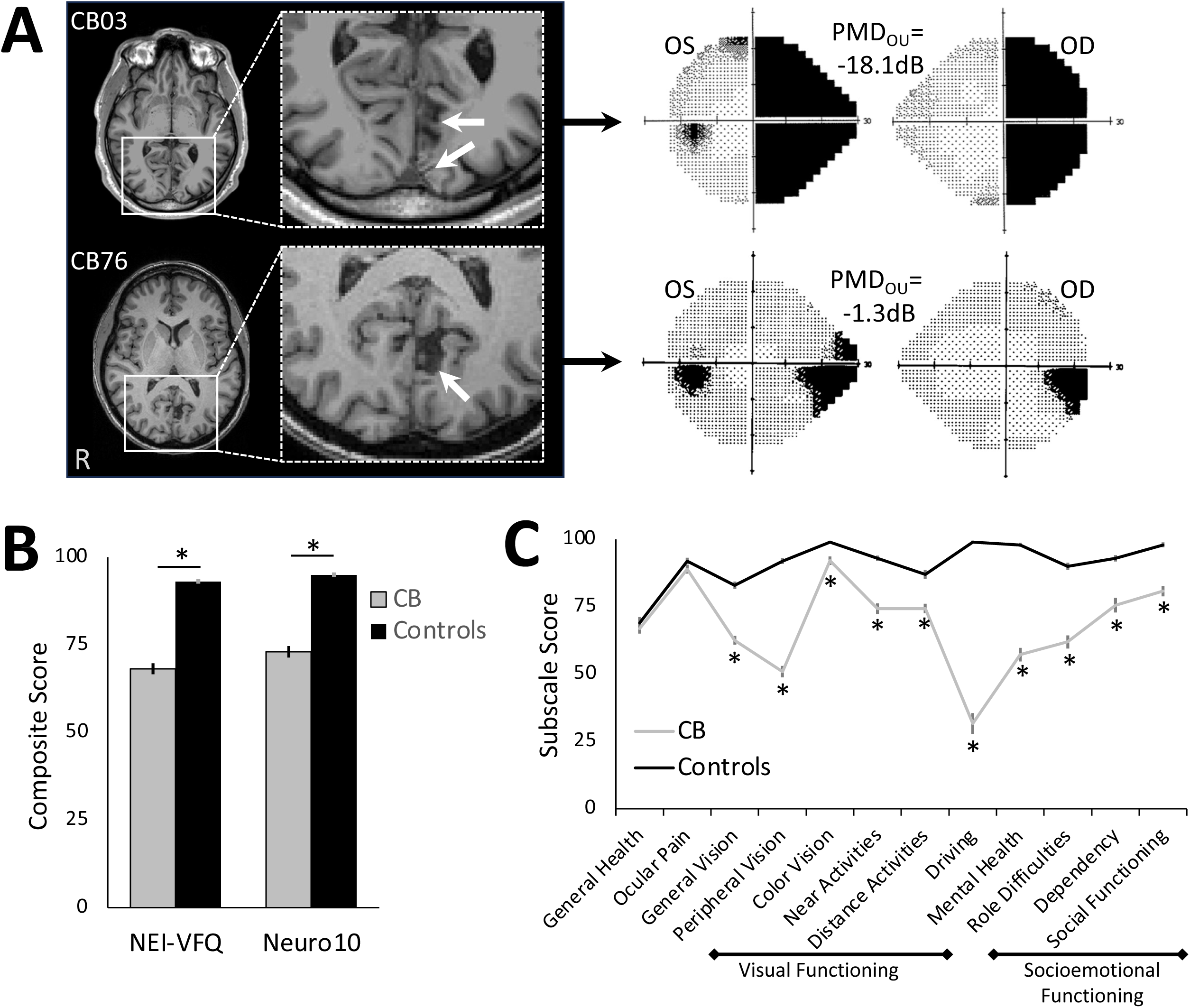
**A**. Sample magnetic resonance images (T1) of 2 CB patients, whose left (OS) and right (OD) eye Humphrey visual fields (HVF) are shown adjacently. For the brain images, radiological convention is used, with the right brain hemisphere (R) on image left. White arrows on enlargements of the regions inside the boxes point to lesion site(s) in the occipital lobe of each patient. On HVFs, black shading denotes a sensitivity of 0 dB, whereas light stippling indicates higher visual sensitivities. Average, binocular (OU) PMDs are indicated for each patient. Note that the larger brain lesion in CB03 gives rise to a larger area of HVF defect and more negative PMD, than the smaller brain lesion in CB76. **B.** Mean NEI-VFQ and Neuro10 composite scores comparing the present cohort of CB patients with previously-published controls (Mangione et al., 2001; Raphael et al., 2006). Mean patient age in our CB cohort did not differ significantly. Controls attained significantly higher composite scores than CB patients on both measures. **C.** Plot of individual NEI-VFQ subscale scores CB patients and the same controls whose composite scores are shown in B. Unsurprisingly, controls scored higher for every subscale except for general health (p=0.480) and ocular pain (p=0.112). Scores evaluating visual functioning and socioemotional functioning are outlined, with “driving” – the most severely affected subscale – separating these two major categories. Error bars in B and C = standard errors of the mean. * p<0.001.

V-QoL surveys were collected prior to and in the absence of training or rehabilitative interventions. CB patients (female/male=26/69) were aged 27-78 (mean±standard deviation=58±12) years old, and ranged from 0.5 to 373.5 (26.0±55.4) months post-stroke (see **Supplemental Table 1** for breakdown per NCT). Patient data were compared to two published reference groups of visually-intact control: one for the National Eye Institute Visual Functioning Questionnaire (NEI-VFQ) and another for its 10-item neuro-ophthalmic supplement (Neuro10). The NEI-VFQ reference group consists of 122 visually-intact participants, 59±14 years old (Mangione et al., 2001); the Neuro10 reference group includes 65 visually-intact controls, 38±12 years old (Raphael et al., 2006). Mean patient age in our CB cohort did not differ significantly from the NEI-VFQ reference group (t_216_=0.56, p=0.578), but the Neuro10 reference group was significantly younger (t_159_=10.38, p<0.0001).

Clinical procedures for the present study were approved by the Western Institutional Review Board for patients enrolled in clinical trial NCT03350919 (WIRB#1181904) and by the Research Subject Review Board at the University of Rochester for NCT04798924 and NCT05098236. All procedures complied with the tenets of the Declaration of Helsinki and were conducted after receiving written, informed consent from each participant.

### Quality of Life Measures

The questionnaires administered included the 25-item version of the NEI-VFQ along with its 14-item appendix, resulting in a total of 39 questions. As per the scoring manual, responses from the NEI-VFQ were used to compute 12 subscale scores, which were averaged together (excepting General Health) to generate a composite score of overall V-QoL (Mangione, 2000). We also administered the 10-item neuro-ophthalmic supplement, which generated an independent Neuro10 composite score. Patients received a paper copy of the survey and were instructed to independently complete the questionnaire at our clinical testing sites. Study staff were present to clarify wording but provided no additional input or assistance.

### Assessment of Visual Field Defect Size and Severity

Relative size and severity of CB visual impairments were estimated using a Humphrey Visual Field Analyzer II-i at two study sites (87 patients), and a Humphrey Visual Field Analyzer 3 at a third site (8 patients), with all sites using a 24-2 testing protocol. Patients were presented with white, Goldman size III stimuli on a white background with a luminance of 11.3 cd/m^2^. Visual sensitivity thresholds for detecting these light targets were calculated using the Swedish Interactive Threshold Algorithm (SITA-Standard). Visual acuity was best-corrected to ≥20/40 using trial lenses, and fixation was controlled with gaze/blind spot automatic settings. Only reliable tests were included in our analysis, defined by fixation loss, false-negative, and false-positive rates <20%. Perimetric mean deviations (PMD), which contrast a participant’s visual field against age-matched, visually intact controls, were calculated monocularly by a proprietary algorithm (Carl Zeiss Meditech), then averaged between eyes to generate a single composite value for visual deficit size/severity in each patient. More negative PMD values indicated greater visual deficit size/severity over the central 54 deg of the visual field.

### Statistical Analyses

Mean NEI-VFQ subscale scores, composite scores, and Neuro10 scores were compared between CB patients and visually-intact controls using unpaired t-tests. Simple linear regressions were used to assess the linear relationships of time post-stroke, age, and PMD with NEI-VFQ and Neuro10 composite scores. Multiple linear regression analyses were used to determine the associations between outcomes — NEI-VFQ subscale scores, NEI-VFQ composite scores, and Neuro10 composite scores — and independent variables (i.e., time post-stroke, PMD, and age) in CB patients in the presence of other risk factors. Analyses were conducted using statistical software (R Studio Version 2023). Statistical significance was set at p<0.05.

## Results

### Characteristics of visual deficits

The present cohort of CB patients presented a wide range of visual deficit sizes and severity, with PMDs ranging from –19.3 to –1.5dB, with a mean of –9.9±4.1dB. Examples of a small and large deficit are shown in **Fig. 1A**. Critically for subsequent analyses, PMD was not significantly correlated with patient age (r^2^=0.047, p=0.113) or time-since-stroke (r^2^=0.062, p=0.065), nor was there a significant correlation between time-since-stroke and patient age (r^2^=0.066, p=0.162).

### Impact of occipital stroke on V-QoL

In CB patients, the mean NEI-VFQ composite score was 68.2±15.3 and the mean Neuro10 score was 73.0±15.8, both significantly lower than control participants, who scored 93.1±6.8 on the NEI-VFQ and 95.0±5.0 on the Neuro10 (**Fig. 1B**; **Table 1**). Individual NEI-VFQ subscales were impaired on 10/12 categories (all but General Health and Ocular Pain; **Table 1**, **Fig. 1C**) in CB patients, but interestingly, none were significantly correlated with PMD (**Supp.** Fig. 1). Differences between CB patients and controls were greatest for Driving, Peripheral Vision, Mental Health, and Role Difficulties (**Table 1**). With respect to driving, 2 CB patients reported that they stopped driving for non-vision related reasons. Per the NEI-VFQ scoring manual, a driving score could not be generated for these patients; therefore, they could not be included in further analysis. Of the remaining CB patients, approximately 53% (49/93) reported giving up driving due to their vision, generating a driving score of zero, while the rest continued to drive.

**Table 1.** Descriptive statistics contrasting NEI-VFQ and Neuro10 scores between CB patients and visually-intact controls. Control data were previously reported controls (Mangione et al., 2001; Raphael et al., 2006). * denotes significance.

| Scale | CB | Control | t | df | p |
| --- | --- | --- | --- | --- | --- |
| NEI-VFQ Composite Score | 68.2±15.3 | 93.1±5.0 | -16 | 216 | < 0.0001* |
| General Health | 66.9±18.5 | 69.0±24.0 | -0.7 | 216 | 0.48 |
| General Vision | 62.6±15.8 | 83.0±15.0 | -9.7 | 216 | < 0.0001* |
| Ocular Pain | 88.9±15.7 | 92.0±13.0 | -1.6 | 216 | 0.112 |
| Near Activities | 74.3±19.1 | 93.0±11.0 | -9.6 | 216 | < 0.0001* |
| Distance Activities | 74.4±16.9 | 87.0±18.0 | -11.6 | 216 | < 0.0001* |
| Social Functioning | 80.8±18.8 | 98.0±10.0 | -8.7 | 216 | < 0.0001* |
| Mental Health | 57.2±24.4 | 98.0±8.0 | -13 | 216 | < 0.0001* |
| Role Difficulties | 61.9±23.7 | 90.0±15.0 | -10.7 | 216 | < 0.0001* |
| Dependency | 75.6±26.3 | 93.0±13.0 | -6.4 | 216 | < 0.0001* |
| Driving | 31.7±37.1 | 99.0±6.0 | -19.7 | 214 | < 0.0001* |
| Color Vision | 92.1±15.2 | 99.0±3.0 | -4.9 | 216 | < 0.0001* |
| Peripheral Vision | 50.9±20.5 | 92.0±12.0 | -18.5 | 216 | < 0.0001* |
| Neuro10 Composite Score | 73.0±15.8 | 95.0±5.0 | -10.9 | 159 | < 0.0001* |

### Relationship Between NEI-VFQ Subscale Scores

We used simple linear regressions to correlate subscale scores with one another. One of the strongest relationships occurred between scores for Distance Activities and Near Activities (**Fig. 2A**). Driving was also significantly correlated with these two subscales (**Fig. 2B, C**), but its strongest correlations were with Mental Health, Role Difficulties, and Dependency (**Fig. 2D-F**). Lastly, scores describing socioemotional functioning (Mental Health, Role Difficulties, Dependency, and Social Functioning) were most strongly intercorrelated with one another. Mental health was most significantly associated with Role Difficulties, Dependency, and Social Functioning (**Fig. 2G-I**).

**Figure 2.**
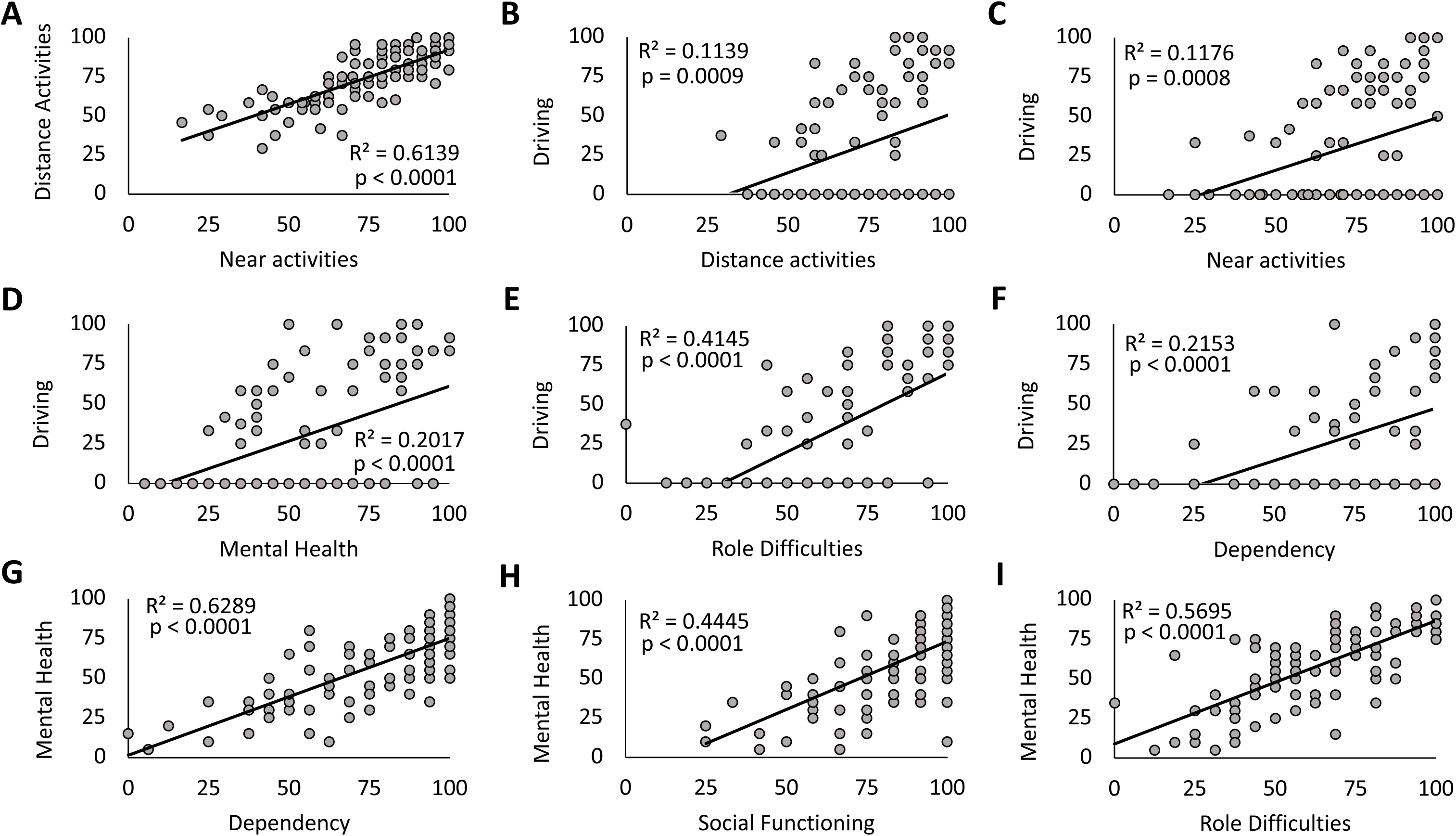
Significant inter-correlations between key visual functioning and socioemotional subscales of the NEI-VFQ. Plots of one subscale score against another with each data point denoting a single CB patient. **A.** Simple regression analysis showed Distance Activities to be correlated significantly and positively with Near Activities. **B.** Similarly, Driving correlated significantly and positively with Distance Activities. **C.** Driving correlated significantly and positively with Near Activities. **D.** Driving correlated significantly and positively with Mental Health. **E.** Driving correlated significantly and positively with Role Difficulties. **F.** Driving correlated significantly and positively with Dependency. **G.** Mental Health correlated significantly and positively with Dependency. **H.** Mental Health correlated significantly and positively with Social Functioning. **I.** Mental Health correlated significantly and positively with Role Difficulties.

### Evolution of QoL scores with time post-stroke

In simple linear regressions, neither PMD nor age were significantly correlated with NEI-VFQ composite scores (**Figs. 3A-B**). PMD was also not correlated with Neuro10 composite scores (**Figs. 3C**), but patient age was (**Fig. 3D**). However, simple linear regressions showed both composite scores to increase with time post-stroke (**Figs. 3E, F**) – correlations that remained significant after adjusting for age and PMD with multiple linear regression analysis (**Table 2**). A one-month increase in time post-stroke increased the average NEI-VFQ and Neuro10 composite scores by ∼0.1 units each in this model (**Table 2**).

**Figure 3.**
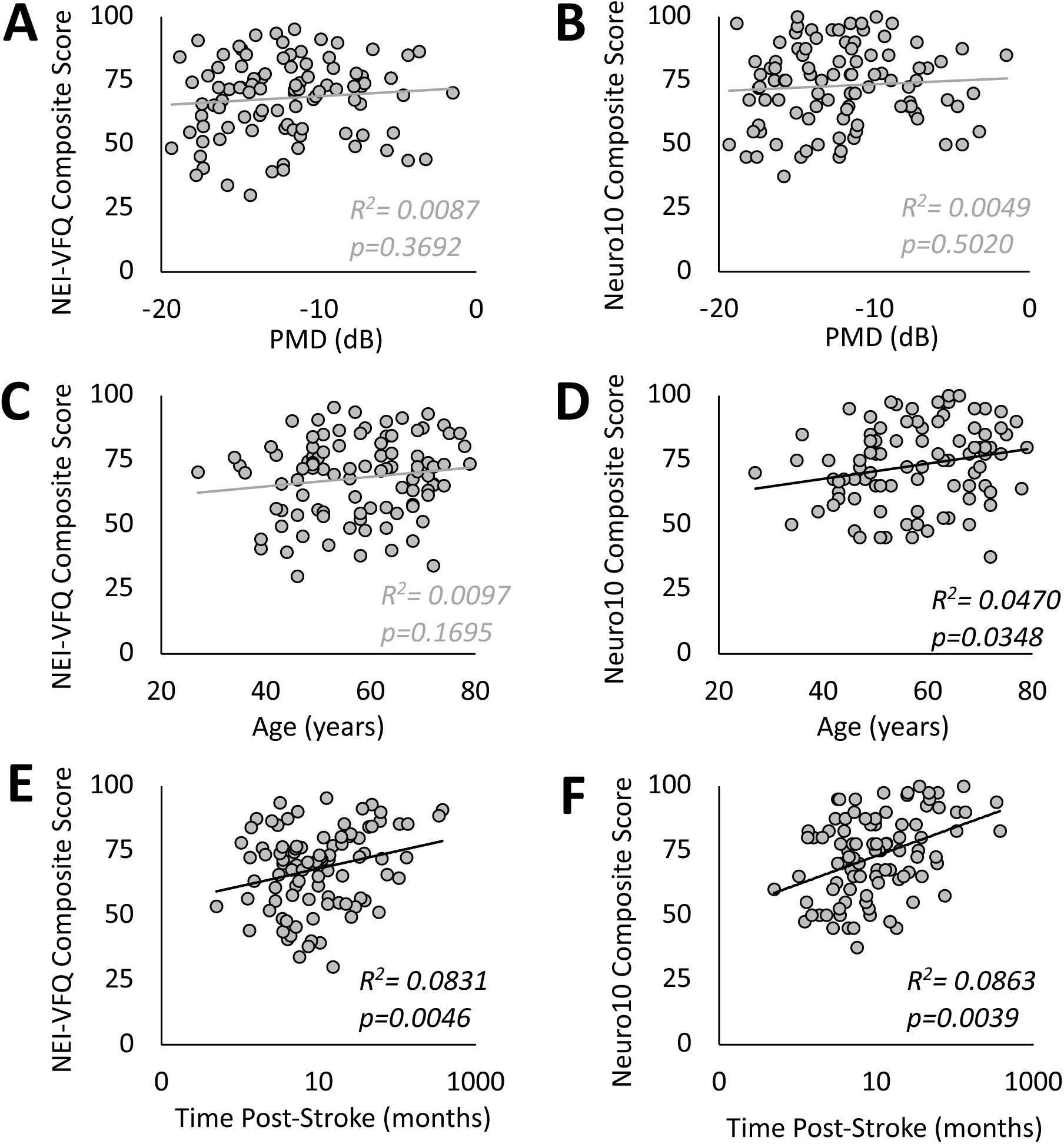
Simple linear regressions correlating age, PMD, and time post stroke individually with NEI-VFQ and Neuro10 composite scores. Plots of composite scores against PMD, age and time post-stroke, with each data point denoting individual patients. PMD was not significantly correlated with NEI-VFQ composite score **(A)** or Neuro10 composite score **(B)**. Age was not significantly correlated with NEI-VFQ composite score **(C)**, but Neuro10 composite score increased significantly with age **(D)**. Both NEI-VFQ composite score **(E)** and Neuro10 score **(F)** increased significantly with time post-stroke – relationships that were maintained after conducting multivariate regression analyses that considered PMD and age (see Table 2 for details).

**Table 2.** Multivariate regression analyses of QoL scores and time post-stroke, adjusted for PMD and age. * indicates significance.

| Scale | Slope | Std Error | t | p |
| --- | --- | --- | --- | --- |
| NEI-VFQ Composite Score | 0.09 | 0.03 | 3.01 | 0.003* |
| General Health | 0.08 | 0.03 | 2.41 | 0.018* |
| General Vision | 0.1 | 0.03 | 3.69 | 0.0004* |
| Ocular Pain | -0.02 | 0.03 | -0.74 | 0.459 |
| Near Activities | 0.06 | 0.04 | 1.59 | 0.116 |
| Distance Activities | 0.08 | 0.03 | 2.5 | 0.014* |
| Social Functioning | 0.04 | 0.03 | 1.28 | 0.203 |
| Mental Health | 0.14 | 0.04 | 3.08 | 0.003* |
| Role Difficulties | 0.11 | 0.04 | 2.5 | 0.014* |
| Dependency | 0.11 | 0.05 | 2.29 | 0.025* |
| Driving | 0.19 | 0.07 | 2.71 | 0.008* |
| Adjusted Driving | 0.11 | 0.05 | 2.21 | 0.033* |
| Color Vision | 0.04 | 0.03 | 1.4 | 0.165 |
| Peripheral Vision | 0.07 | 0.04 | 1.88 | 0.064 |
| Neuro10 Composite Score | 0.08 | 0.03 | 2.94 | 0.004* |

Two participants were enrolled >300 months post-stroke: their enrollment dates post-stroke were each more than 300 months longer than the mean enrollment time post-stroke (26.0 months) and both more than 325 months longer than the median enrollment time post-stroke (9.1 months). To assess whether these two data points influenced the slope estimate in our linear regression models, i.e., whether they acted as statistical leverage points, we repeated the multivariate regression excluding these two patients; both the NEI-VFQ and Neuro10 composite scores still increased significantly with time post-stroke (p=0.034 and p=0.0008, respectively).

Finally, adjusting for PMD and age, 7/12 NEI-VFQ subscale scores improved with increasing time post-stroke: General Health, General Vision, Distance Activities, Mental Health, Role Difficulties, Dependency, and Driving (**Table 2**), even when the analysis excluded participants who stopped driving post-stroke (“adjusted driving” score, **Table 2**). The remaining 5 subscale scores did not change significantly with time since stroke: Ocular Pain, Near Activities, Social Functioning, Color Vision, and Peripheral Vision.

## Discussion

The present cross-sectional analysis of post-occipital stroke patients adds to the growing discussion regarding the impact of visual field deficits on vision-related quality of life. Ours is one of the first studies to include both subacute (<6 months post-stroke) and chronic (>6 months post-stroke) patients in the same analysis, while focusing solely on unilateral stroke sustained in adulthood. This allowed us to assess a wider range of times post-stroke than previous work. Moreover, not binning patients by deficit size or lesion age (Gall et al., 2010) allowed novel correlations to emerge between these parameters and V-QoL. Specifically, we uncovered that V-QoL appears to increase with time post-stroke, irrespective of deficit size or patient age. Subscale analyses provided key insights into the likely drivers of this unexpected relationship.

### Occipital stroke reduces vision-related QoL

We compared V-QoL scores among our sample of CB patients to two reference groups of visually-intact controls — one for the NEI-VFQ and another for the Neuro10 supplement. The NEI-VFQ control group is referenced extensively in studies quantifying the impact of ophthalmic diseases on V-QoL (Cahill, Banks, Stinnett, & Toth, 2005; Clemons et al., 2008; Hariprasad et al., 2008; Ma et al., 2002; Schiffman, Jacobsen, & Whitcup, 2001). This group was well-matched in age to our sample of CB patients, who nonetheless had significantly lower NEI-VFQ composite scores. CB patients also scored lower than controls on the Neuro10 supplement, although the reference group for the latter was younger than our CB patients. Overall, reductions in both NEI-VFQ and Neuro10 composite scores confirmed that V-QoL was impaired in our cohort of CB stroke-only patients.

Notably, 24-2 binocular Humphrey PMD, an objective proxy for central visual deficit size and severity, failed to correlate with NEI-VFQ or Neuro10 composite scores when adjusting for age and time post-stroke. Subscale analyses revealed a similar lack of correlation between PMD and all NEI-VFQ subscales (**Supp.** Fig. 1), suggesting that CB patients’ perceived difficulty in visual and socioemotional functioning is not related to the objective severity or size of their central visual deficit. This finding contrasts somewhat with prior literature reporting improved V-QoL scores with increased central visual field sparing (Gall et al., 2010; Papageorgiou et al., 2007), and greater improvement in V-QoL with greater spontaneous improvement in deficit size up to 6 months post-stroke (Tharaldsen et al., 2020). Our results may instead reflect a greater contribution of other social functioning factors on V-QoL scores in the present cohort, or be due to our multivariate analyses, which accounted for both age and time since stroke.

One important consideration in the context of the present study is that while 24-2 HVF perimetry is the most commonly-used clinical tool for quantifying visual deficits in CB, it does not capture the entirety of the visual field, and may thus miss some of the deficit. In future studies, quantification and categorization (hemianopia, scotoma, degree of macular sparing, etc.) of the CB deficit with whole-field methods (e.g., Goldmann or automated kinetic perimetry) may address this problem and it is conceivable that they may yield stronger correlations with V-QoL than Humphrey perimetry. Finally, V-QoL in this patient population could be impacted by patient visual acuity. Presently, all patients were best-corrected to 20/40 or better, and foveal acuity is not thought to be impacted in homonymous hemianopia, which typically presents with normal foveal sensitivity (as was the case in all of our patients’ HVFs). Finally, previous reports found no difference in V-QoL in occipital stroke patients when comparing patients with good vs poor central acuity (Gall et al., 2010).

### Subscale-specific impairments

When investigating the impact of occipital stroke on subscales of the NEI-VFQ, CB patients scored significantly lower than age-matched controls for each subscale except for General Health and Ocular Pain, consistent with prior work (Mangione et al., 2001). The lack of impairment in these two subscales was unsurprising, as occipital stroke is not associated with physical discomfort to the eye, and CB patients enrolled met stringent medical and functional criteria to participate in the studies analyzed (see Methods). In fact, our CB self-reported General Health scores were as good as those of healthy controls and suggest that differences in other subscales were due to the stroke’s impact on vision, instead of other impairments seen in the general stroke population.

The observed impairments among CB patients for subscales describing visual ability (General Vision, Peripheral Vision, Near Activities, and Distance Activities) were expected, since homonymous hemianopia is characterized by deficits that often span large segments of the visual periphery. Impairments in socioemotional subscales (Mental Health, Role Difficulties, Dependency, and Social Functioning) may instead be related to post-stroke depression, which has been documented in up to half of sufferers within the first 5 years post-stroke (Ayerbe, Ayis, Crichton, Wolfe, & Rudd, 2014; Hackett & Pickles, 2014; Kim & Choi-Kwon, 2000). Impairments in Mental Health and related socioemotional subscales may also be associated with the reduced ability to drive. Driving was the subscale where CB patients scored most poorly compared to controls, with 53% reporting that they gave up driving entirely due to their eyesight. Driving cessation has also been associated with increased depressive symptoms in a general population of older adults (Ragland, Satariano, & MacLeod, 2005). This was confirmed here, with Driving scores most strongly correlated with Mental Health, Role Difficulties and Dependency scores, and driving, or lack thereof, may also be a determining factor in role fulfillment.

On-road testing found CB patients to have noticeable, but not intractable, driving deficits. Chronic CB patients have at-fault accident rates 2.6 times higher than the visually-intact population, motivating driving restrictions for hemianopic patients (McGwin, Wood, Huisingh, & Owsley, 2016). However, on-road testing found that up to 82% of drivers with homonymous hemianopia committed no obvious errors while driving (Elgin et al., 2010), and when evaluators were masked to the participants’ condition, 73% of hemianopic and 88% of quadrantopic patients were deemed fit to drive (Wood et al., 2009). Given the large impact of driving on quality of life, evaluation for driving safety and rehabilitation may offer a simple and cost-effective method of improving CB patients’ quality of life. Finally, we note that pass/fail results for on-road testing and simulator environments show good fidelity in CB patients (Ungewiss et al., 2018), suggesting that simulated tests could providing a good-fidelity, safe, naturalistic space for examination of CB driving ability (Bowers, 2016; Bowers, Mandel, Goldstein, & Peli, 2009).

### V-QoL increases naturally with time since stroke

A key finding here was that V-QoL increases with time post-stroke, even when adjusting for age and PMD. Our multivariate analysis included a much larger range of post-stroke times (<1 month to over 31 years) than any prior work (Gall et al., 2010; Gall et al., 2009; Papageorgiou et al., 2007). Improvement in health-related QoL with time has been previously reported in the general stroke population, with occipital stroke patients showing slower recovery than their visually-intact counterparts (Gall et al., 2010). A distinct possibility is that this slower rate of improvement is a feature of global QoL for CB patients, resulting from the lack of available rehabilitation compared to that for speech or motor impairments. Nonetheless, the gradual improvement in V-QoL over time offers hope for CB patients dealing with life changes in the early stages post-stroke.

To understand what aspects of visual functioning drove the observed larger V-QoL at later times post-stroke, we examined individual NEI-VFQ subscale scores, adjusting for age and PMD. Seven improved significantly over time: General Health, General Vision, Distance Activities, Mental Health, Role Difficulties, Dependency, and Driving. It is notable that these scores were significantly correlated with each other in our subscale analyses. As such, any of these categories may drive the observed *overall* improvement, enhancing other categories in the process. Alternatively, they may improve independently, or with some mixed amount of driving effects from specific subscales. The remaining five subscales (Ocular Pain, Near Activities, Social Functioning, Color Vision, and Peripheral Vision) did not improve over time post stroke. Ocular Pain was not impacted by this stroke, leaving Near Activities, Social Functioning, Color Vision and Peripheral Vision as potential targets for rehabilitation and counseling.

In our earlier comparison with visually intact controls, we noted that Mental Health and Role Difficulties were among the most impaired subscales in CB patients, likely reflecting a loss of self-sufficiency and professional ability. Increased scores for Mental Health, Role Difficulties, and Dependency over time may reflect the gradual development of lifestyle adjustments that help patients contend with their visual impairment, and recover some self-sufficiency and professional functioning. Alternatively, patients may simply become accustomed to their deficit and “learn to live with it”, despite the burdens it imposes.

Finally, higher V-QoL scores at later times post-stroke do not negate the fact that as a cohort, CB patients remained impaired relative to controls in all subscales excepting Ocular Pain and General Health (**Fig. 1**). Thus, accelerating recovery or enhancing its magnitude may be critical for ultimately improving patient outcomes.

### Limitations

A key limitation of the present study was an inability to assess V-QoL in the same patient, as it progressed over time. Instead, we relied upon the different times post-stroke across individuals to make inferences about V-QoL natural history. It is hoped that future studies will address this problem and rule out the possibility of a sampling bias within the CB population who volunteers for research studies such as ours, long after their stroke.

We should also note that differences in age may have confounded comparison of Neuro10 scores between CB patients and controls, since the reference group for those scores was significantly younger than our CB patients (Raphael et al., 2006). However, given the extreme degree of V-QoL impairment in CB evidenced by the NEI-VFQ, and its lack of correlation with age, it is likely our observations would hold for the Neuro10, even with appropriately-matched controls.

Finally, although the NEI-VFQ has been used extensively to quantify V-QoL in CB patients, we note that it was originally designed for patients with ophthalmic disease (Gall et al., 2009). As such, it may not robustly capture all aspects of V-QoL that pertain to CB. Development and validation of a CB-specific questionnaire could significantly improve our understanding of this condition and its impact on daily life.

### Conclusions and importance of knowledge gained

V-QoL is significantly reduced by occipital strokes sustained in adulthood. Neither deficit size/severity nor age were significant predictors of this reduction. However, our cross-sectional design and large range of times post-stroke revealed that both composite scores and some key, individual sub-scores to be greater at later *versus* earlier times post-stroke. This suggests a spontaneous increase in V-QoL with time, even after adjusting for patient age and deficit severity. These results provide new, potentially key information for patients, clinicians and researchers as to the main drivers of V-QoL after occipital stroke. It is now possible to describe what changes patients might expect as they progress in their stroke recovery. Just as importantly, it alerts the field to the fact that V-QoL may improve spontaneously after occipital stroke, a trend against which vision restoration therapies in this patient population should be measured in order to assess their true impact.

## Acknowledgements

The authors would like to thank Adin Reisner, Lisa Blanchard, Chrys Callan for their assistance in recruiting and screening patients for the studies reported. We also thank Terrance Schaefer and Rachel Hollar for performing the HVF testing on these patients and Dr. Brent Johnson for advice on statistical analyses.

## Data Availability

The data that support the findings of this study are available from the corresponding author upon reasonable request.

## Author Contacts

Neil Dogra –

Bryan V. Redmond –

Selena Lilley –

Brent A. Johnson –

Byron L. Lam –

Madhura Tamhankar –

Steven E. Feldon –

Berkeley Fahrenthold –

Jingyi Yang –

Krystel R. Huxlin –

Matthew R. Cavanaugh –

## References

1. Ayerbe, L., Ayis, S. A., Crichton, S., Wolfe, C. D., & Rudd, A. G. (2014). Natural history, predictors and associated outcomes of anxiety up to 10 years after stroke: the South London Stroke Register. Age Ageing, 43(4), 542–547. doi:10.1093/ageing/aft208

2. Bowers, A. R. (2016). Driving with homonymous visual field loss: a review of the literature. Clinical & experimental optometry, 99(5), 402–418. 10.1111/cxo.12425

3. Bowers, A. R., Mandel, A. J., Goldstein, R. B., & Peli, E. (2009). Driving with hemianopia, I: Detection performance in a driving simulator. Invest Ophthalmol Vis Sci, 50(11), 5137–5147. doi:10.1167/iovs.09-3799

4. Cahill, M. T., Banks, A. D., Stinnett, S. S., & Toth, C. A. (2005). Vision-related quality of life in patients with bilateral severe age-related macular degeneration. Ophthalmology, 112(1), 152–158. doi:10.1016/j.ophtha.2004.06.036

5. Chen, C. S., Lee, A. W., Clarke, G., Hayes, A., George, S., Vincent, R., Crotty, M. (2009). Vision-Related Quality of Life in Patients with Complete Homonymous Hemianopia Post Stroke. Topics in Stroke Rehabilitation, 16(6), 445–453. doi:10.1310/tsr1606-445

6. Clemons, T. E., Gillies, M. C., Chew, E. Y., Bird, A. C., Peto, T., Figueroa, M., Macular Telangiectasia Research, G. (2008). The National Eye Institute Visual Function Questionnaire in the Macular Telangiectasia (MacTel) Project. Invest Ophthalmol Vis Sci, 49(10), 4340–4346. doi:10.1167/iovs.08-1749

7. Dombovy, M. L., Sandok, B. A., & Basford, J. R. (1986). Rehabilitation for stroke: a review. Stroke, 17(3), 363–369. doi:10.1161/01.str.17.3.363

8. Elgin, J., McGwin, G., Wood, J. M., Vaphiades, M. S., Braswell, R. A., DeCarlo, D. K., Owsley, C. (2010). Evalution of On-Road Driving in Persons with Hemianopia and Quadrantanopia. Am J Occup Ther, 64(2), 268–278. doi:10.5014/ajot.64.2.268

9. Gall, C., Franke, G. H., & Sabel, B. A. (2010). Vision-related quality of life in first stroke patients with homonymous visual field defects. Health and Quality of Life Outcomes, 8(1), 33. doi:10.1186/1477-7525-8-33

10. Gall, C., Lucklum, J., Sabel, B. A., & Franke, G. H. (2009). Vision– and health-related quality of life in patients with visual field loss after postchiasmatic lesions. Invest Ophthalmol Vis Sci, 50(6), 2765–2776. doi:10.1167/iovs.08-2519

11. George, S., Hayes, A., Chen, C., & Crotty, M. (2011). Are vision-specific quality of life questionnaires important in assessing rehabilitation for patients with hemianopia post stroke? Top Stroke Rehabil, 18(4), 394–401. doi:10.1310/tsr1804-394

12. Gilhotra, J. S., Mitchell, P., Healey, P. R., Cumming, R. G., & Currie, J. (2002). Homonymous visual field defects and stroke in an older population. Stroke, 33(10), 2417–2420. doi:10.1161/01.str.0000037647.10414.d2

13. Gray, C. S., French, J. M., Bates, D., Cartlidge, N. E., Venables, G. S., & James, O. F. (1989). Recovery of visual fields in acute stroke: homonymous hemianopia associated with adverse prognosis. Age Ageing, 18(6), 419–421. doi:10.1093/ageing/18.6.419

14. Hackett, M. L., & Pickles, K. (2014). Part I: frequency of depression after stroke: an updated systematic review and meta-analysis of observational studies. Int J Stroke, 9(8), 1017–1025. doi:10.1111/ijs.12357

15. Hariprasad, S. M., Mieler, W. F., Grassi, M., Green, J. L., Jager, R. D., & Miller, L. (2008). Vision-related quality of life in patients with diabetic macular oedema. Br J Ophthalmol, 92(1), 89–92. doi:10.1136/bjo.2007.122416

16. Horton, J. C. (2005a). Disappointing results from Nova Vision’s visual restoration therapy. Br J Ophthalmol, 89(1), 1–2. doi:10.1136/bjo.2004.058214

17. Horton, J. C. (2005b). Vision restoration therapy: confounded by eye movements. Br J Ophthalmol, 89(7), 792–794. doi:10.1136/bjo.2005.072967

18. Jones, S. A., & Shinton, R. A. (2006). Improving outcome in stroke patients with visual problems. Age Ageing, 35(6), 560–565. doi:10.1093/ageing/afl074

19. Jongbloed, L. (1986). Prediction of function after stroke: a critical review. Stroke, 17(4), 765–776. doi:10.1161/01.str.17.4.765

20. Kim, J. S., & Choi-Kwon, S. (2000). Poststroke depression and emotional incontinence: correlation with lesion location. Neurology, 54(9), 1805–1810. doi:10.1212/wnl.54.9.1805

21. Liu, K. P. Y., Hanly, J., Fahey, P., Fong, S. S. M., & Bye, R. (2019). A Systematic Review and Meta-Analysis of Rehabilitative Interventions for Unilateral Spatial Neglect and Hemianopia Poststroke From 2006 Through 2016. Archives of Physical Medicine and Rehabilitation, 100(5), 956–979. 10.1016/j.apmr.2018.05.037

22. Ma, S. L., Shea, J. A., Galetta, S. L., Jacobs, D. A., Markowitz, C. E., Maguire, M. G., & Balcer, L. J. (2002). Self-reported visual dysfunction in multiple sclerosis: new data from the VFQ-25 and development of an MS-specific vision questionnaire. Am J Ophthalmol, 133(5), 686–692. doi:10.1016/s0002-9394(02)01337-5

23. Mangione, C. M. (2000). The National Eye Institute 25-Item Visual Functioning Questionnaire (VFQ-25).

24. Mangione, C. M., Lee, P. P., Gutierrez, P. R., Spritzer, K., Berry, S., & Hays, R. D. (2001). Development of the 25-item National Eye Institute Visual Function Questionnaire. Arch Ophthalmol, 119(7), 1050–1058. doi:10.1001/archopht.119.7.1050

25. Mangione, C. M., Lee, P. P., Pitts, J., Gutierrez, P., Berry, S., & Hays, R. D. (1998). Psychometric properties of the National Eye Institute Visual Function Questionnaire (NEI-VFQ). NEI-VFQ Field Test Investigators. Arch Ophthalmol, 116(11), 1496–1504. doi:10.1001/archopht.116.11.1496

26. McGwin, G., Wood, J. M., Huisingh, C., & Owsley, C. (2016). Motor Vehicle Collision Involvement among Persons with Hemianopia and Quadrantanopia. Geriatrics, 1(19). doi:10.3390/geriatrics1030019

27. Melnick, M. D., Tadin, D., & Huxlin, K. R. (2016). Relearning to See in Cortical Blindness. The Neuroscientist, 22(2), 199–212. doi:10.1177/1073858415621035

28. Papageorgiou, E., Hardiess, G., Schaeffel, F., Wiethoelter, H., Karnath, H. O., Mallot, H., Schiefer, U. (2007). Assessment of vision-related quality of life in patients with homonymous visual field defects. Graefes Arch Clin Exp Ophthalmol, 245(12), 1749–1758. doi:10.1007/s00417-007-0644-z

29. Plant, G. T. (2005). A work out for hemianopia. Br J Ophthalmol, 89(1), 2. doi:10.1136/bjo.2004.053173

30. Pollock, A., Hazelton, C., Henderson, C. A., Angilley, J., Dhillon, B., Langhorne, P., Shahani, U. (2012). Interventions for visual field defects in patients with stroke. Stroke, 43(4), e37–e38. doi:10.1161/strokeaha.111.639815

31. Pollock, A., Hazelton, C., Henderson, C. A., Angilley, J., Dhillon, B., Langhorne, P., Shahani, U. (2011). Interventions for visual field defects in patients with stroke. Cochrane Database Syst Rev (10), CD008388. doi:10.1002/14651858.CD008388.pub2

32. Pollock, A., Hazelton, C., Rowe, F. J., Jonuscheit, S., Kernohan, A., Angilley, J., Campbell, P. (2019). Interventions for visual field defects in people with stroke. Cochrane Database Syst Rev, 5(5), CD008388. doi:10.1002/14651858.CD008388.pub3

33. Ragland, D. R., Satariano, W. A., & MacLeod, K. E. (2005). Driving cessation and increased depressive symptoms. J Gerontol A Biol Sci Med Sci, 60(3), 399–403. doi:10.1093/gerona/60.3.399

34. Raphael, B. A., Galetta, K. M., Jacobs, D. A., Markowitz, C. E., Liu, G. T., Nano-Schiavi, M. L., Balcer, L. J. (2006). Validation and test characteristics of a 10-item neuro-ophthalmic supplement to the NEI-VFQ-25. Am J Ophthalmol, 142(6), 1026–1035. doi:10.1016/j.ajo.2006.06.060

35. Reinhard, J., Schreiber, A., Schiefer, U., Kasten, E., Sabel, B. A., Kenkel, S., Trauzettel-Klosinski, S. (2005). Does visual restitution training change absolute homonymous visual field defects? A fundus controlled study. Br J Ophthalmol, 89(1), 30–35. doi:10.1136/bjo.2003.040543

36. Rowe, F. J. (2013). Care provision and unmet need for post stroke visual impairment Final Report.

37. Rowe, F. J., Hepworth, L. R., Conroy, E. J., Rainford, N. E. A., Bedson, E., Drummond, A., Sackley, C. (2019). Visual Function Questionnaire as an outcome measure for homonymous hemianopia: subscales and supplementary questions, analysis from the VISION trial. Eye (Lond), 33(9), 1485–1493. doi:10.1038/s41433-019-0441-z

38. Sahraie, A. (2007). Induced visual sensitivity changes in chronic hemianopia. Current Opinion in Neurobiology, 20, 661–666.

39. Saionz, E. L., Busza, A., & Huxlin, K. R. (2022). Rehabilitation of visual perception in cortical blindness. Handb Clin Neurol, 184, 357–373. doi:10.1016/B978-0-12-819410-2.00030-8

40. Saionz, E. L., Feldon, S. E., & Huxlin, K. R. (2020). Rehabilitation of cortically-induced visual field loss. Current Opinion in Neurology, 34, 67–74.

41. Schiffman, R. M., Jacobsen, G., & Whitcup, S. M. (2001). Visual functioning and general health status in patients with uveitis. Arch Ophthalmol, 119(6), 841–849. doi:10.1001/archopht.119.6.841

42. Tharaldsen, A. R., Sand, K. M., Dalen, I., Wilhelmsen, G., Naess, H., Midelfart, A., Group, t. N.-O. R. (2020). Vision-related quality of life in patients with occipital stroke. Acta Neurol Scand, 141, 509–518.

43. Ungewiss, J., Kübler, T., Sippel, K., Aehling, K., Heister, M., Rosenstiel, W., Papageorgiou, E. (2018). Agreement of driving simulator and on-road driving performance in patients with binocular visual field loss. Graefe’s Archive for Clinical and Experimental Ophthalmology, 256(12), 2429–2435. doi:10.1007/s00417-018-4148-9

44. Wood, J. M., McGwin, G., Elgin, J., Vaphiades, M. S., Braswell, R. A., DeCarlo, D. K., Owsley, C. (2009). On-Road Driving Performance by Persons with Hemianopia and Quadrantanopia. Invest Ophthalmol Vis Sci., 50(2), 577–585. doi:10.1167/iovs.08-2506

45. Zhang, X., Kedar, S., Lynn, M. J., Newman, N. J., & Biousse, V. (2006). Natural history of homonymous hemianopia. Neurology, 66(6), 901–905. doi:10.1212/01.wnl.0000203338.54323.22

