## Supplemental Materials for "The Natural History of Vision-Related Quality of Life after Unilateral Occipital Stroke"

| Clinical Trial Identifier | # F/M | Age (years) | Time post-stroke (months) | Binocular PMD (dB) | NEI-VFQ Composite Score | Neuro10 Composite Score |
|---|---|---|---|---|---|---|
| NCT04798924 | 10/15 | 50.8±11.7 | 3.5±1.3 | -11.3±4.2 | 69.1±14.2 | 68.4±13.8 |
| NCT03350919 | 10/32 | 59.5±9.9 | 45.1±76.7 | -13.1±4.0 | 67.1±17.4 | 75.5±18.0 |
| NCT05098236 | 6/22 | 61.1±11.9 | 17.4±25.2 | -11.3±3.8 | 68.9±12.0 | 73.4±13.2 |

**Supplementary Table 1. Demographic characteristics across the three NCT analyzed in the present study.** Other than female (F) and male (M) numbers, all data are reported as mean ± SD.

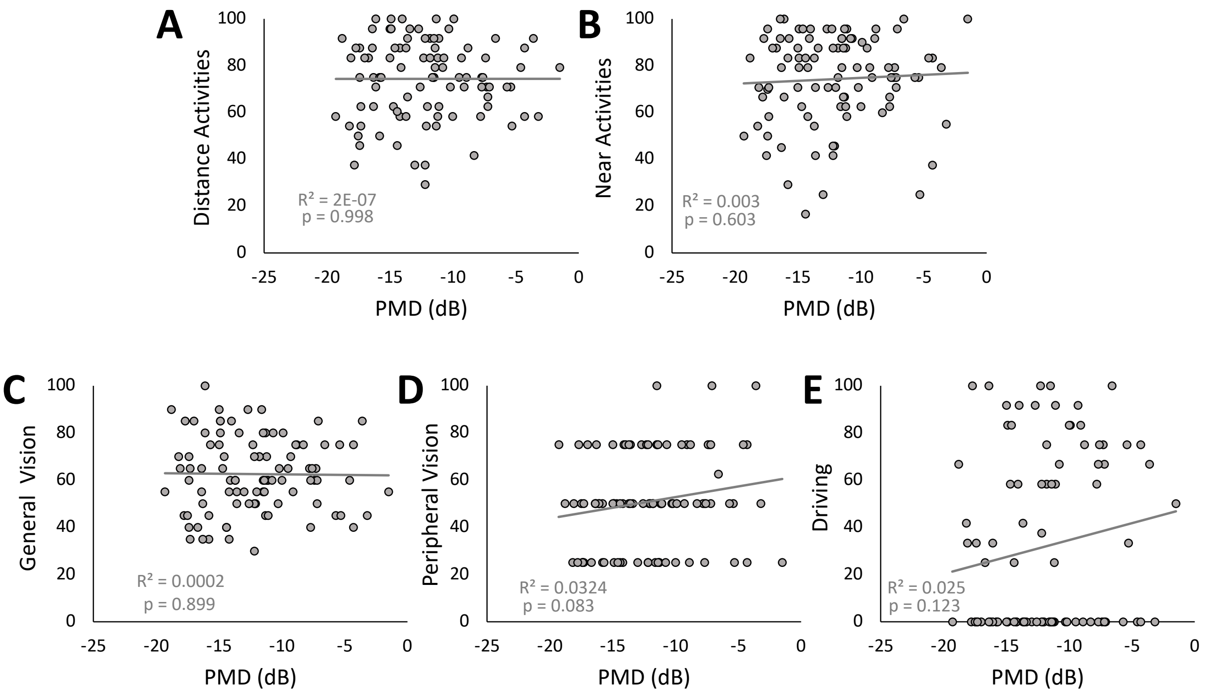


**Supplementary Figure 1.** Simple linear regressions correlating PMD with key subscales describing visual functioning. **A**, PMD was not significantly correlated with scores for distance activities. **B**, PMD was not significantly correlated with scores for near activities. **C**, PMD was not significantly correlated with scores for general vision. **D**, PMD was not significantly correlated with scores for peripheral vision. **E**, PMD was not significantly correlated with scores for driving.
